# Now casting and Forecasting of COVID-19 outbreak in the National Capital Region of Delhi

**DOI:** 10.1101/2020.05.01.20087783

**Authors:** Bharathnag Nagappa, Manikandanesan Sakthivel, Yamini Marimuthu, Aayushi Rastogi, Archana Ramalingam, Shiv Kumar Sarin

## Abstract

**Objectives:** The study aimed to estimate the disease burden due to COVID-19 in the scenarios of unchecked spread and with various public health interventions in New Delhi.

**Methods:** We adopted Susceptible, Exposed, Infected and Recovered (SEIR) model to estimate the course of COVID-19 outbreak in Delhi population and effect of public health intervention on the pandemic. We first estimated the basic reproductive rate (R_0_) based on the evidence from Wuhan, then ran the model considering no intervention implemented, followed by case isolation, social distancing, and lockdown, each implemented in isolation and in combinations to estimate the number of cases. Markov’s model was used to estimate the number of cases in various clinical scenarios of the disease. Sensitivity analysis conducted to estimate the effect of asymptomatic cases on case based interventions.

**Results:** Estimated R_0_ in Delhi population was 6.18 (range 4.15 – 12.2). Effective reproductive rate (R_t_) was least for case isolation (3.5). Lockdown showed highest reduction (28%) in number of prevalent cases on peak day and 22% reduction in patients in need of intensive care unit (ICU). Case isolation and lockdown together resulted in 50% reduction in number of prevalent cases and 42% reduction in patients in need of ICU care. Sensitivity analysis showed that the effect of case isolation was inversely proportionate to the proportion of asymptomatic (hidden) cases.

**Conclusions:** Interventions should be implemented in combinations of individual and community level interventions to gain better outcome. Identifying and isolation of all cases as early as possible is important to flatten the pandemic curve.

## Introduction

Clusters of individuals with pneumonia and clinical presentation similar to severe acute respiratory syndrome coronavirus (SARS-CoV) reported from Wuhan in December, 2019.^[1,2]^ On January 12, 2020, a novel strain of coronavirus (severe acute respiratory syndrome corona virus 2 (SARS-CoV-2)) was isolated from these clusters of pneumonia cases and the new respiratory illness named as coronavirus disease 2019 (COVID-19).^[3]^ Evidence suggesting that the Huanan seafood wholesale market was the initial transmission site for the novel virus.^[4]^ The market was closed and disinfection measures were taken to prevent further transmission.^[5]^ Since then Cases of COVID-19 have been reported in health-care workers and family members of cases with 67,794 cases and 3,805 deaths confirmed in Hubei province as of March 15, 2020.^[4,6,7]^ Later the centre of pandemic shifted to Italy and then to the United states which currently accounts for the highest number of cases in the world. Older individuals (aged >60 years) and people with chronic health conditions are reported to be more susceptible to severe disease.^[6]^ Rapid spread of the virus led the Chinese Government to restrict movement in affected cities, with the cessation of public transport and cancellation of flights.^[8,9]^ Despite extensive efforts to prevent onward spread, 203 countries and territories outside the mainland of China have reported imported cases. Internationally, ongoing local transmission of SARS-CoV-2 has been confirmed in 149 countries and territories.^[7]^ In India, 10,197 tested positive, 1343 cured and 392 deaths related COVID-19 were reported as on 15^th^ April, 2020.^[10]^ India has reported local transmission and may enter stage three of pandemic soon. India had taken measures like, home quarantining all international travellers, national level lockdown, promoting social distancing, tracing and quarantining of contacts of all positive cases to prevent the further spread of diseases.^[11]^ Even with all these measures India is still noticing the increase in COVID-19 cases. Cases were reported among health care workers, family members and other close contacts of COVID-19 cases. There might be an exponential increase in COVID-19 cases in India. Because of asymptomatic cases, under reporting and misdiagnosis, it is difficult for policy makers to understand the exact burden of disease and act accordingly.

Maharasthra followed by the National Capital Region of Delhi, account for the largest number of reported cases in the country today.^[10]^ Modelling studies are needed to understand the disease burden and potential impact of early interventions to guide the policy makers in this early stage of COVID-19 in India.

## Material and Methods

### Epidemiological Model

We used Susceptible (S), Exposed (E), Infected (I), Recovered (R) (SEIR) model to estimate the disease burden in the community. Where ‘S’ represents the number of susceptible persons, ‘E’ represents the total number of persons in latent period, ‘I’ represents the total infected and symptomatic individuals and ‘R’ represents the total people recovered from the disease at the given time (t).^[12]^ This model gives insights on the flow of people from susceptible to exposed, exposed to infected and infected to recovered phase. The model considered the relationship between basic reproductive rate (R_0_) and total infectious period to estimate the transmissibility coefficient. To estimate the contact probability, we divided the susceptible population by total population. As the infectious period equals to inverse of recovery rate, the model considered one over infectious period to estimate the recovery rate.^[13]^ SEIR model assumes that the population are in closed compartments.^[13]^ But, in the real scenario, many imported cases (with history of travel outside India and tested positive) were reported from Delhi. To adjust for it, we added the imported cases on the reported day into the total infected cases till 14^th^ day of stopping international flights. The SEIR is model given below.^[14]^

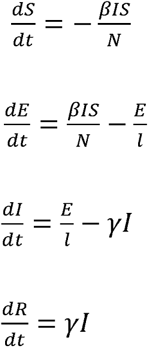

We used Markov’s probability model to estimate the mild disease, severe disease, number of patients needing intensive care unit (ICU) admission and deaths due to COVID-19.^[15]^ We applied daily recovery rate separately for severe and mild diseases, to account for the difference in time to recover from the disease in mild and severe cases.

To estimate the basic reproductive rate of Delhi, we multiplied the population density ratio of Delhi and area where R_0_ was taken from.^[14]^

R_0_ in Delhi population = (Population density of Delhi/population density of Wuhan)* R_0_ in Wuhan population

To estimate the effective reproductive rate (R_t_) when interventions are implemented, we multiplied the reduction fractions (rf) assumed for different interventions (Table I) and basic reproductive rate. Community-level interventions are given to all individuals irrespective of diseases status, but individual-level interventions (cases isolation, contacts isolation or quarantine) are limited to cases and their close contacts. Community-level interventions decrease the transmission and prevent the complete spectrum of diseases whereas individual-level interventions decrease the transmission from only symptomatic/identified cases and would not alter the transmission from asymptomatic/ hidden cases. Therefore, while estimating the number of cases, effect of community level interventions were applied to both asymptomatic and symptomatic cases and effect of individual level interventions were applied only to symptomatic cases. In multiple interventions scenario, effect of community level interventions on R_t_ was estimated first, and effect of individual level intervention applied to R_t_ of community level intervention.

**Table I:** Contribution to case pool and effect of interventions on transmission among contact groups of COVID-19 cases.

|  |  |  |  |  |  |  |
| --- | --- | --- | --- | --- | --- | --- |
| Illness acquired at | Proportion contributed to case pool<sup>[24]</sup> | Reduction achieved by intervention |  |  |  |  |
|  |  | Social distancing | Lock down | case isolation | case isolation + Lock down | case isolation + Social distancing |
| Household contact | 48.7 | 0 | 0 | 25 | 25 | 25 |
| Workplace and hubs contact | 30.5 | 0 | 90 | 60 | 96 | 60 |
| Community contact | 20.8 | 25 | 25 | 60 | 70 | 70 |
| Total reduction achieved |  | 5.2 | 32.65 | 42.95 | 56.01 | 45.03 |
| With the current response from the population we assumed at least 50% shows the compliance to social distancing. |  |  |  |  |  |  |

R_t_ in single intervention scenario

R_t_ = rf * R_0_

R_t_ in Multiple intervention scenario

R_t_ = (rf_ci_ + (1-rf_ci_)*rf_i_)* R_0_

R_t_ – effective reproductive rate

R_0_ – Basic reproductive rate

rf – reduction fraction

rf_ci_ – reduction fraction due to community level intervention

rf_i_ – reduction fraction due to individual level intervention

### COVID-19 infection parameters and assumptions

According to 2011 census data, Delhi population was 1.64 crore.^[16]^ Using birth and death rate of we estimated the current Delhi population to be 1.9 crore.^[17]^ In the SEIR model, we assumed that no individual had the immunity against COVID-19 in the study setting. We assumed 2^nd^ March 2020 as day zero, when the first index case reported. There was no data available on parameters of COVID-19 disease in Indian community; hence we used the data from international studies. The parameters included were incubation period (5 days)^[4,18,19]^ of COVID-19, time needed for recovery from disease (7 days for Mild disease and 15 days for severe disease), and time of death from the onset of symptoms (18 days).^[18]^ Studies reported that the average infectious period was 10 days^[20]^ with 30% of the cases showing no symptoms.^[21]^ As per the estimates given in literatures, around 80% of symptomatic persons experience mild disease, 20% experience severe disease; 5% need admission in ICU and 3.8% of them may die due to COVID-19.^[18,22]^

Existing literature gave basic reproductive rate in broad range. Rocklöv J et al estimated the basic reproductive rate to be 14.8, before taking any control measure in Diamond Princes cruise ship.^[14]^ Studies from other parts of Wuhan were given basic reproductive rate in range of 2.2 to 6.47 with mean of 3.28.^[23]^ Based on these evidences, we estimated the basic reproductive rate for Delhi by adjusting for the difference in the population density.

We classified the contacts of cases into household contacts, contacts in hubs (contacts in workplace and public gatherings), and community contacts (neighbours, contacts during travelling and other outdoor activities). We used estimates of SARS-Cov 2 to obtain the proportion of cases resulted from each type of contact [Table I].^[24]^ We considered all interventions, government of Delhi implemented and isolation of both case and contacts to estimate the burden of COVID-19 in Delhi. In social distancing intervention, we assume at least 50% of the individuals have followed the intervention in community and its effect is negligible in workplace and house due to repeated contact and closed environment. We considered 4 days delay in case isolation^[4,25]^ due to delay in health care seeking (2 days), sample collection, transport (1 day), testing and reporting (1 day). To estimate the reduction fraction in case isolation we subtracted the days of delay in case isolation from total infectious period, assuming equal probability of transmission of the virus throughout the infectious period in workplace and community contact. In lockdown intervention (closure of workplace except for essential services, educational institutes, recreational sites, banning of public gathering, stopping of transportation except for essential services), we considered all people stay at home, thus, have nil effect on house hold contacts, But reduce 90% of workplace transmission (since 10% may occur during provision of essential services) and community transmission similar to social distancing (people continued maintain contact in closed communities) [Table I].

**Table II:** Estimated basic reproductive rate and effective reproductive rate of COVID-19 in Delhi population.

| Parameters |  | Estimates |
| --- | --- | --- |
| Mean basic reproductive rate |  | 6.18 |
| Minimum basic reproductive rate |  | 4.15 |
| Maximum basic reproductive rate |  | 12.2 |
| Effective reproductive rate | Social distancing | 5.86 |
|  | Lockdown | 4.16 |
|  | Case isolation | 3.56 |
|  | Case isolation and social distancing | 3.40 |
|  | Case isolation and lock down | 2.72 |
|  | Case and contact isolation | 1.62 |

In Delhi, case isolation was implemented from day zero, social distancing was implemented from day 15 and lockdown was implemented from day 20 after reporting of the index case.^[26,27]^ For social distancing and lockdown we assumed that the effect of intervention may be shown after 7 days (half of maximum incubation period) of implementing intervention.

### Sensitivity analysis

We have done sensitivity analysis for variation in the proportion of asymptomatic cases. For the purpose of sensitivity analysis, variation in proportion of asymptomatic cases was adopted from study conducted by Nishiura H et al.^[21]^ Variation in asymptomatic cases affects the estimation when case targeted interventions implemented. Therefore we conducted sensitivity analysis for variations in asymptomatic cases when scenarios included case isolation intervention.

Ethics: There are no ethical concerns related to the study since all the data are taken from the official public domain from the respective institutions.

## Results

In the study, we estimated the R_0_ first. Mean R_0_ for Delhi population was estimated to be 6.18 and it ranges from 4.15 to 12.2. We also estimated the effective reproductive rate (R_t_) for different intervention. Least R_t_ was seen when isolation of cases and contacts are strictly implemented followed by lockdown combined with case isolation [Table II].

We estimated the effect of interventions social distancing, lockdown and case isolation separately as well as in combination as they were implemented by the government of Delhi. The study estimated that the outbreak will reach its peak in 94 days, if no interventions are implemented. The peak of the epidemic curve can be delayed by 4 days if only social distancing was implemented, by 35 days when only lockdown was implemented, by 31 days when only case isolation is implemented. The same will delay by 35 days when case isolation along with social distancing was implemented, by 28 days when case isolation with 21 days lockdown was implemented and by 69 days when case isolation along with continued lockdown is implemented. When individual interventions were considered, lockdown had the maximum effect on pandemic [Figure 1]. Lockdown alone reduced the maximum number of prevalent cases on peak day, mean of prevalent cases per day, and mean number of patients needed ICU per day by 28%, 22% and 22%, respectively, by reducing the transmission at workplace and community contacts [Figure 3].

**Figure 1:**
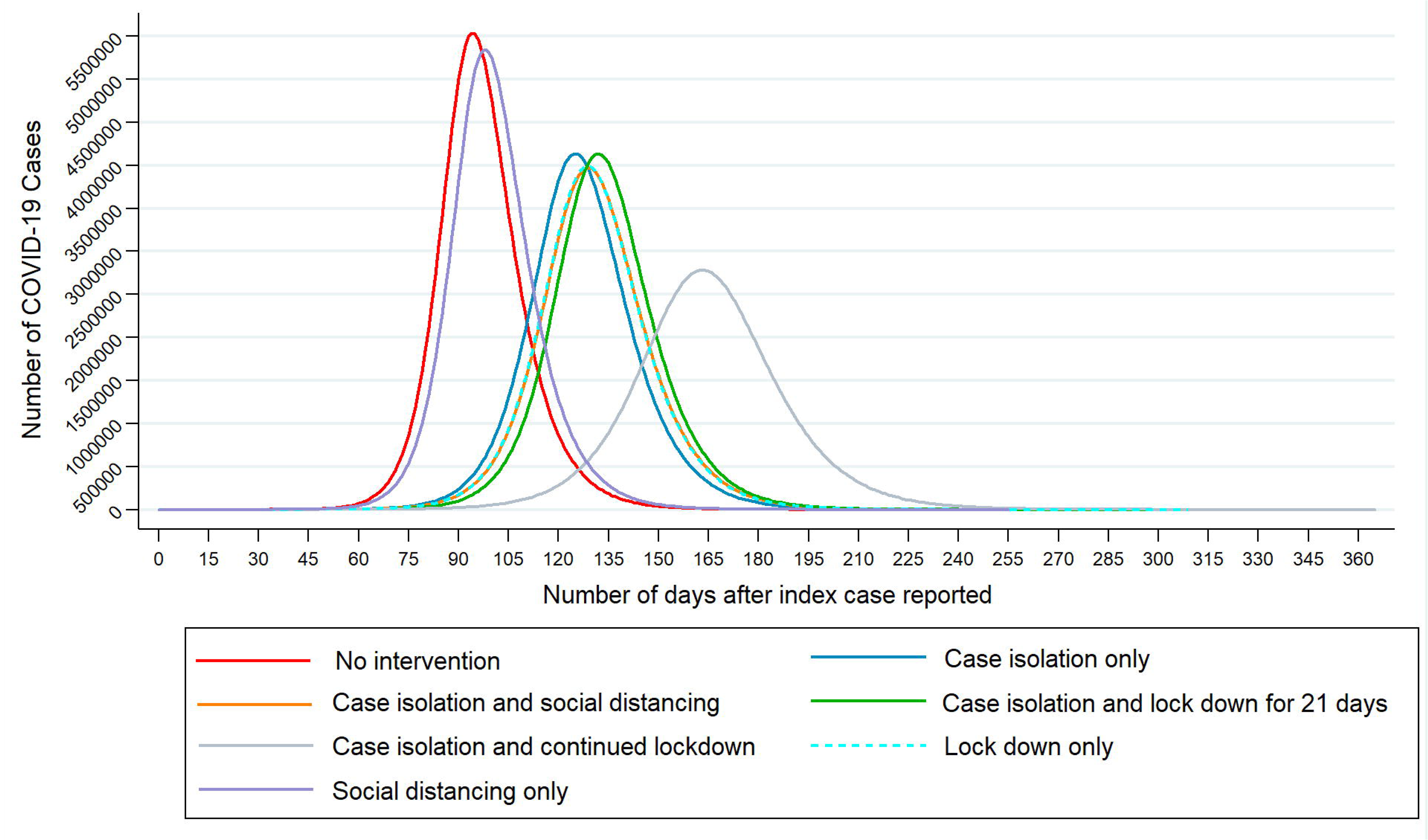
Estimation of COVID-19 cases using SEIR model in various public health interventions.

**Figure 2:**
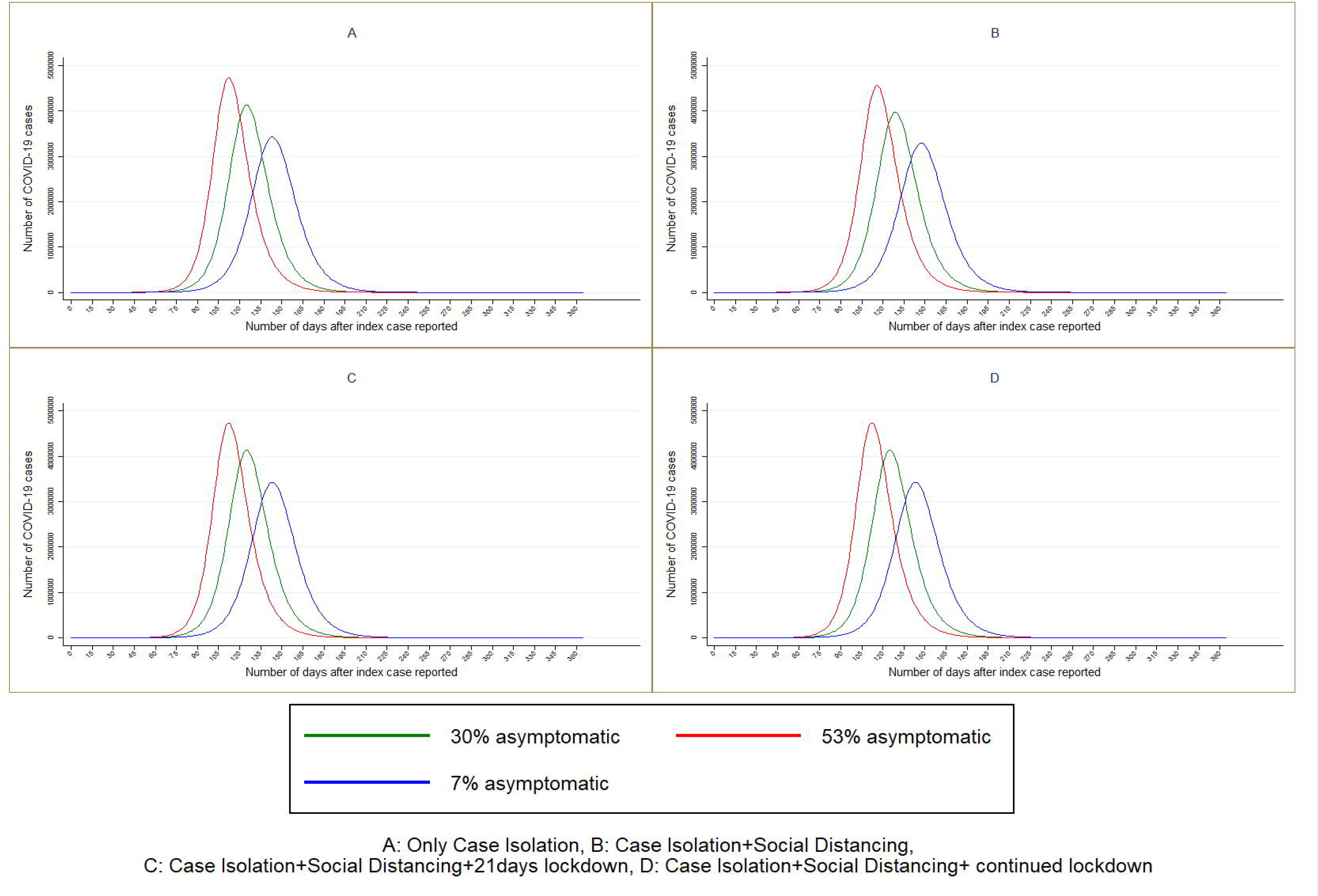
Sensitivity analysis for proportion of asymptomatic cases.

**Figure 3:**
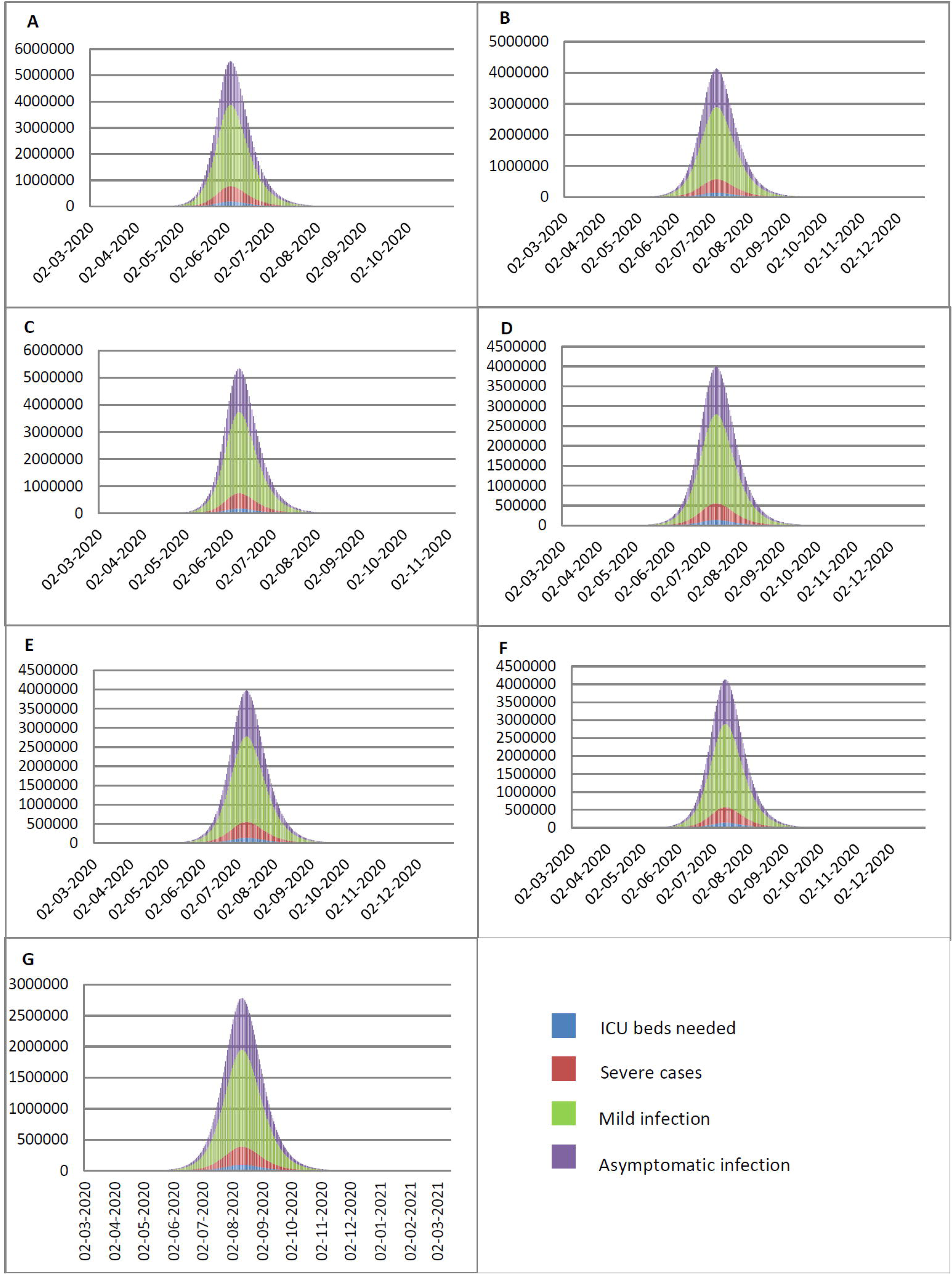
(A) No intervention (B) Only case isolation (C) Social distancing only (D) Lock down only (E) Case isolation and Social distancing (F) Case isolation, Social distancing and Lock down for 21 days (G) Case isolation, Social distancing and Continued lockdown

It was observed that various interventions implemented in combination would delay the peak of pandemic and flatten the pandemic curve. In the intervention of case isolation with social distancing, peak of pandemic occurs 3 days earlier as compared to case isolation with lockdown for 21 days. But reduction in number of prevalent cases per day and reduction in mean of number of patients needed ICU admission was higher in case isolation with social distancing (28% and 22%, respectively) compared to case isolation with lockdown for 21 days (25% and 22%, respectively). Case isolation with continued lockdown reduced the number of prevalent cases on peak day by 50% and showed mean reduction of ICU beds by 42%. Details of the effect of various public health interventions on COVID-19 outbreak are given in table III.

**Table III:** Effect of public health interventions on COVID-19 pandemic in Delhi.

|  |  |  |  |  |  |  |  |
| --- | --- | --- | --- | --- | --- | --- | --- |
| Interventions | no intervention | Social distancing | Lock down | case isolation | case isolation with social distancing | case isolation with 21 day lock down | Case isolation with continued lock down |
| Days to reach Peak of pandemic | 94 | 98 | 129 | 125 | 129 | 132 | 163 |
| Maximum number of prevalent cases/day (on peak day) | 5538364 | 5347571 | 3993430 | 4136277 | 3972078 | 4136017 | 2783885 |
| reduction in Maximum number of prevalent cases/day (on peak day) |  | 3 | 28 | 25 | 28 | 25 | 50 |
| Mean of prevalent cases/day | 626525 | 610373 | 487840 | 501683 | 485857 | 490354 | 363151 |
| reduction in mean of prevalent cases |  | 3 | 22 | 20 | 22 | 22 | 42 |
| maximum number of severe cases/day | 581528 | 561495 | 419310 | 434309 | 417068 | 434282 | 292308 |
| Mean of sever cases/day | 65785 | 64089 | 51223 | 52677 | 51015 | 51487 | 38131 |
| Maximum number of patients needed ICU admission | 193843 | 187165 | 139770 | 144770 | 139023 | 144761 | 97436 |
| Reduction in Maximum number of patients needed ICU admission in single day |  | 3 | 28 | 25 | 28 | 25 | 50 |
| mean of patients needed ICU | 21928 | 21363 | 17074 | 17559 | 17005 | 17162 | 12710 |
| Reduction in ICU needed |  | 3 | 22 | 20 | 22 | 22 | 42 |

The study also estimated the number of asymptomatic cases, symptomatic cases with mild infection, severe infection and patients in need of ICU care, which are shown in figure 3 of supplementary material. It is estimated that around 193,843 patients need ICU care on peak day of pandemic which can be reduced 97436 if we implement case isolation and continue the lockdown. Figure 4 shows the comparison of actual number of confirmed COVID-19 cases in Delhi with the estimated numbers till 30.04.2020. Sensitivity analysis showed that if asymptomatic (hidden) cases are less, all individual level interventions showed maximum effect [Figure 2]. When proportion of asymptomatic cases was 7%, case isolation delayed the peak of pandemic by 53 days with 41% reduction in prevalent cases on peak day and case isolation with continued lockdown delayed the peak by 93 days with 60% reduction in prevalent cases. When proportion of asymptomatic cases was 53%, case isolation alone and in combination with continued lockdown, delayed the pandemic by 22 days with 18% reduction in prevalent cases on peak day and by 52 days with 41% reduction in prevalent cases, respectively.

**Figure 4:**
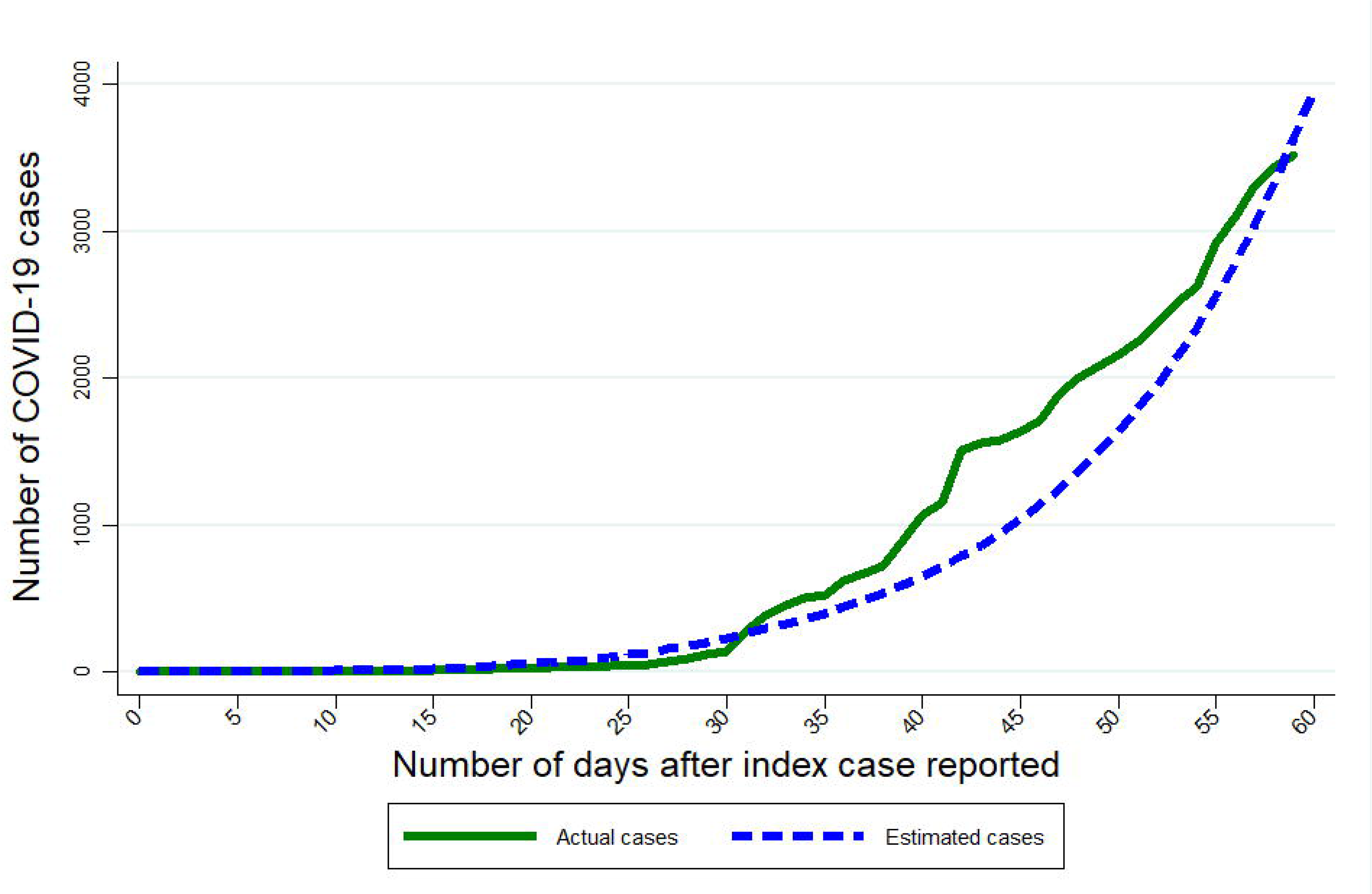
Comparison between actual and estimated cases in Delhi till 30.04.2020.

## Discussion

The results of our study forecast the possible course of the pandemic in the National Capital Region of Delhi and the likely effect of various public health interventions considered by the Government of India and the Government of Delhi. Accordingly estimates shows that pandemic may reach its peak on 94^th^ day with around 55 lakh prevalent cases on peak day, if no public health interventions implemented. By case isolation and lockdown we could delay the pandemic by 69 days with around 28 lakh (50% reduction) prevalent cases on the peak day.

Before forecasting the course of pandemic, we estimated the basic reproductive rate (R_0_) for Delhi to be 6.18 (range 4.15-12.2).^[23]^ Based on data obtained from diamond prince ship, R_0_ was estimated to be 14.7.^[14]^ The difference in estimated R_0_ might be due to the variation in population density. Population density of Delhi was 11320 whereas the population density of Diamond Prince Ship was 24400.^[14,16]^ If we adjust to population density then R_0_ 14.7 in cruise ship is equivalent to R_0_ 6.86 in Delhi, which is comparable to the estimates in our study. However, the model has not considered the difference in contact rate among people, which may lead to slight variations in the R_0_. We preferred the R_0_ estimate based on evidence from Wuhan for forecasting the pandemic in Delhi, as these estimates were based on data obtained before any large scale community interventions were implemented.

Our study estimated that the outbreak will reach its peak in 94 days, if no interventions are implemented. When interventions implemented separately, lockdown showed maximum effect with delaying the pandemic by 35 days and when implemented in combination case isolation with continued lockdown showed the maximum effect. Lockdown alone was able to reduce number prevalent cases on peak day of pandemic by 28%, where case isolation alone reduced by 25% and social distancing alone reduced by 3%. These findings showed that lockdown had the maximum effect on the transmission of disease. Case isolation showed the maximum effect on reproductive rate in our estimations. These differences were mainly due to the effect of asymptomatic cases. Currently, majority of the countries are testing only the symptomatic cases and the same is applicable to Delhi also. The asymptomatic cases who will not be identified and isolated will continue to transmit the disease in the community.^[25]^ Therefore the effect of case isolation decreased due to asymptomatic cases. Lockdown is a community-level intervention which is applied to everyone irrespective of their disease status. This will decrease the transmission from both asymptomatic and symptomatic cases. We incorporated these points into the model while estimating the number of prevalent cases. In sensitivity analysis, it was shown that when the proportion of asymptomatic cases decreased, the effect of case isolation increased. To decrease the number of hidden cases and case isolation to be successful, we need to increase the diagnostic testing capacity of health system for COVID-19. These findings also highlight that due to high proportion of asymptomatic cases and limited capacity of health system to trace and test all contacts of positive cases; case-based interventions are having limited scope in flattening epidemic curve. To have a better outcome we need to combine both case-based and community-based approach. These findings are further supported by evidences from European countries, compiled by imperial collage COVID-19 response team.^[28]^ The study conducted by Koo et al also showed that combined interventions including case based approach and community level interventions were more successful in flattening the pandemic curve.^[29]^

The findings suggests that, even though lockdown showing better results compared to other interventions, implementing it for short duration would not show the better results. Case isolation with lockdown for 21 days delayed peak of pandemic by 3 more days compared to case isolation combined with social distancing but still showed higher number of prevalent cases on peak day. To have better outcome lockdown should be continued for longer duration. If it implemented for short duration, loss may outweighs the benefits of the intervention.

The strength of SEIR model is that, it is a tested model to estimate the size of outbreak with local transmission. It gives good estimates of outbreak in a defined population, which helps in planning and policy making to control the outbreak in the population. It also provides us the rate of increase or decrease in number of cases which is very useful to understand the dynamics of outbreak. The model considers that the disease starts from single foci and spreads to the rest of the population. In current scenario, we have multiple imported cases which could have affected the results. However, we added the imported cases (who had travel history and diagnosed to be COVID-19 positive in Delhi) on the day of reporting to infected pool.

The present model also has a few limitations. The populations living in a rapidly developing new metropolis may be highly heterogeneous due to caste, religion or economic power, and if that affects the basic reproductive rate in significant scale, the estimates might vary. However, the NCR is a well-developed and rather stable metropolis and we believe it is relatively less heterogeneous in nature than many two tier towns in the country and thus will have lesser effect on the estimates.^[16]^ Literatures were uncertain about when the infected person starts to shed the virus, and the most commonly found time is 12 hours before symptoms appear.^[20]^ We had assumed that at the end of incubation period, infected person starts to shed the virus which applied for both symptomatic and asymptomatic cases.

In summary, interventions implemented in Delhi are time buying interventions to prepare and act to mitigate the effect of pandemic and to make it manageable. Interventions should be implemented in combinations of individual level and community level interventions to gain better outcome. Identifying and isolation of all cases as early as possible is important to flatten the pandemic curve.

## Data Availability

Data will be made accessible if required.

## Acknowledgement

NilFinancial support: Nil

Conflict of Interest: None declared

